# Renin-angiotensin system blockers and mortality in COVID-19: a territory-wide study from Hong Kong

**DOI:** 10.1101/2020.12.21.20248645

**Authors:** Jiandong Zhou, Gary Tse, Sharen Lee, Wing Tak Wong, Xingsong Wang, William KK Wu, Tong Liu, Zhidong Cao, Daniel Dajun Zeng, Ian Chi Kei Wong, Bernard Man Yung Cheung, Qingpeng Zhang

## Abstract

**Aims:** Renin–angiotensin system blockers such as angiotensin-converting enzyme inhibitors (ACEIs) and angiotensin receptor blockers (ARBs) may increase the risk of adverse outcomes in COVID-19. In this study, the relationships between ACEI/ARB use and COVID-19 related mortality were examined.

**Methods:** Consecutive patients diagnosed with COVID-19 by RT-PCR at the Hong Kong Hospital Authority between 1^st^ January and 28^th^ July 2020 were included.

**Results:** This study included 2774 patients. The mortality rate of the COVID-19 positive group was 1.5% (n=42). Those who died had a higher median age (82.3[76.5-89.5] vs. 42.9[28.2-59.5] years old; P<0.0001), more likely to have baseline comorbidities of cardiovascular disease, diabetes mellitus, hypertension, and chronic kidney disease (P<0.0001). They were more frequently prescribed ACEI/ARBs at baseline, and steroids, lopinavir/ritonavir, ribavirin and hydroxychloroquine during admission (P<0.0001). They also had a higher white cell count, higher neutrophil count, lower platelet count, prolonged prothrombin time and activated partial thromboplastin time, higher D-dimer, troponin, lactate dehydrogenase, creatinine, alanine transaminase, aspartate transaminase and alkaline phosphatase (P<0.0001). Multivariate Cox regression showed that age, cardiovascular disease, renal disease, diabetes mellitus, the use of ACEIs/ARBs and diuretics, and various laboratory tests remained significant predictors of mortality.

**Conclusions:** We report that an association between ACEIs/ARBs with COVID-19 related mortality even after adjusting for cardiovascular and other comorbidities, as well as medication use. Patients with greater comorbidity burden and laboratory markers reflecting deranged clotting, renal and liver function, and increased tissue inflammation, and ACEI/ARB use have a higher mortality risk.

**Key Points:**

- We report that an association between ACEIs/ARBs with COVID-19 related mortality even after adjusting for cardiovascular and other comorbidities, as well as medication use.
- Patients with greater comorbidity burden and laboratory markers reflecting deranged clotting, renal and liver function, and increased tissue inflammation, and ACEI/ARB use have a higher mortality risk.

## Introduction

The use of angiotensin□converting enzyme (ACE) inhibitors and angiotensin receptor blockers (ARBs) has been associated with poor disease outcomes in coronavirus disease 2019 (COVID-19). For example, a population-based cohort study from Korea found that ACE-I or ARB therapy in patients with severe COVID-19 was associated with the occurrence of severe complications and increased in-hospital mortality [1]. By contrast, other studies did not identify an increased risk of severe COVID-19 [2, 3] or mortality [4] among patients taking ACEIs or ARBs. A systematic review and meta-analysis of 33 studies found that ACE inhibitor use was marker of increased mortality risk in some, but not all, COVID-19 disease settings [5], whilst another found that The risk of mortality and severe outcomes are also unchanged among COVID-19 patients taking ACEI/ARB [6]. Nevertheless, prior use of RAAS inhibitors was associated with lower risk mortality from COVID-19 specifically in patients with hypertension [7]. Given these conflicting findings, we examined whether the use of ACEIs/ARBs is related to COVID-19 mortality using territory-wide data from Hong Kong.

## Methods

### Study design and population

This study was approved by the Institutional Review Board of the University of Hong Kong/Hospital Authority Hong Kong West Cluster. This was a retrospective, territory-wide cohort study of patients undergoing COVID-19 RT-PCR testing between 1^st^ January and 28^th^ July 2020. The patients were identified from the Clinical Data Analysis and Reporting System (CDARS), a territory-wide database that centralizes patient information from individual local hospitals to establish comprehensive medical data, including clinical characteristics, disease diagnosis, laboratory results, and drug treatment details. The system has been previously used by both our team and other teams in Hong Kong [8-11]. The list of conditions identified is detailed in the **Supplementary Appendix**. Patients and the public were not engaged in this study.

### Outcomes and statistical analysis

All-cause mortality was the primary outcome for the COVID-19 positive group, with last date of mortality status on 8^th^ August 2020. Continuous variables were presented as median (95% confidence interval [CI] or interquartile range[IQR]) and categorical variables were presented as count (%). The Mann-Whitney U test was used to compare continuous variables. The χ^2^ test with Yates’ correction was used for 2×2 contingency data. Statistical analyses were performed using RStudio software (Version: 1.1.456) and Python (Version: 3.6).

## Results

A total of 2774 patients with confirmed COVID-19 were included. Their baseline characteristics are shown in **Table 1**. A total of 42 deaths occurred (1.5%) in the 2774 patients who were diagnosed with COVID-19. Those who died had a higher median age (82.3[76.5-89.5] vs. 42.9[28.2-59.5] years old), more likely to have baseline comorbidities of cardiovascular disease, diabetes mellitus, hypertension, and chronic kidney disease. Moreover, they were more frequently prescribed ACEI/ARBs at baseline, and steroids, lopinavir/ritonavir, ribavirin and hydroxychloroquine during admission. Finally, patients who died had a higher white cell count, higher neutrophil count, lower platelet count, prolonged PT and activated partial thromboplastin time, higher D-dimer, troponin and lactate dehydrogenase. Moreover, creatinine, alanine transaminase, aspartate transaminase and alkaline phosphatase were significantly higher for the mortality group.

**Table 1.**
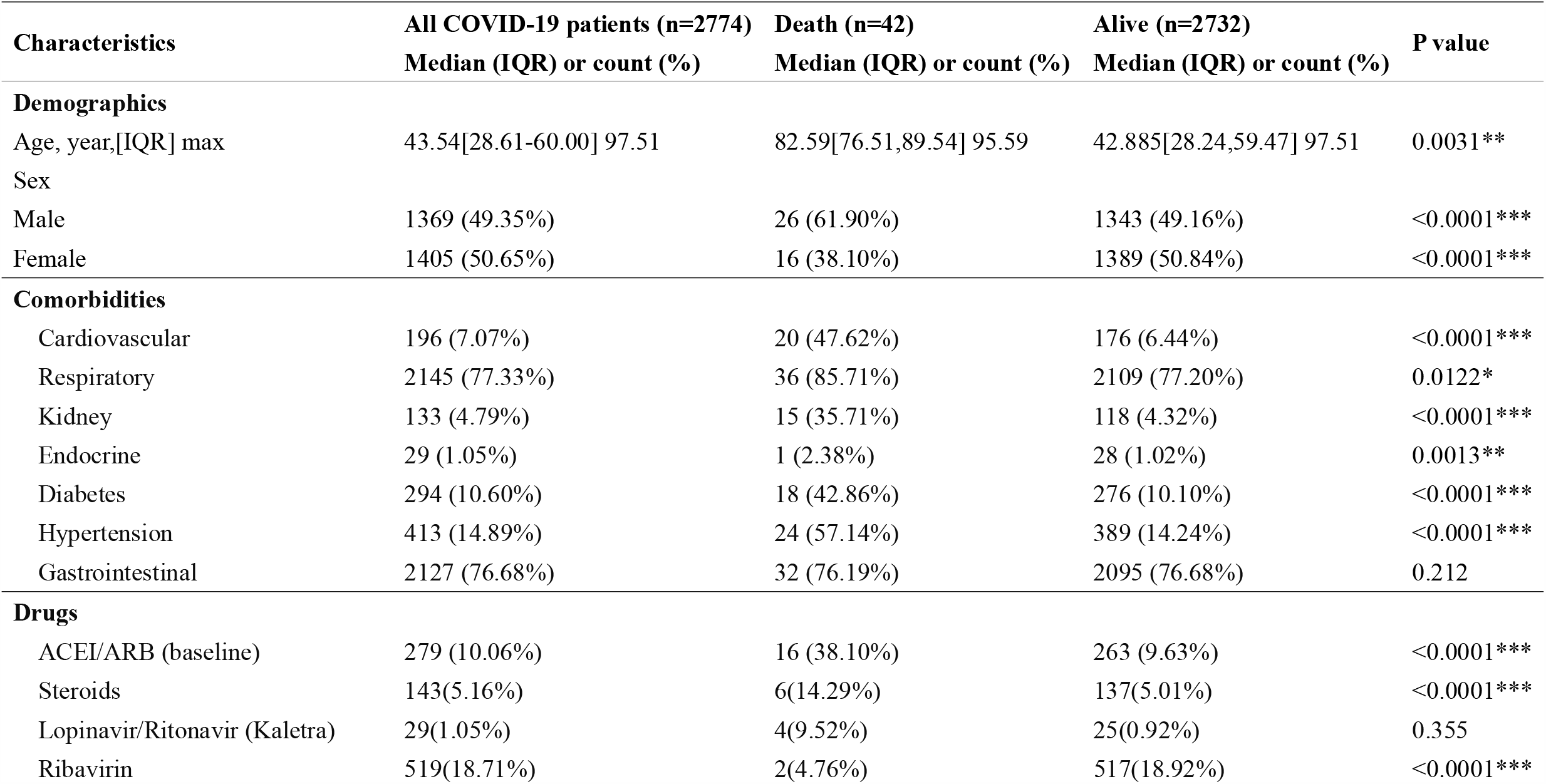
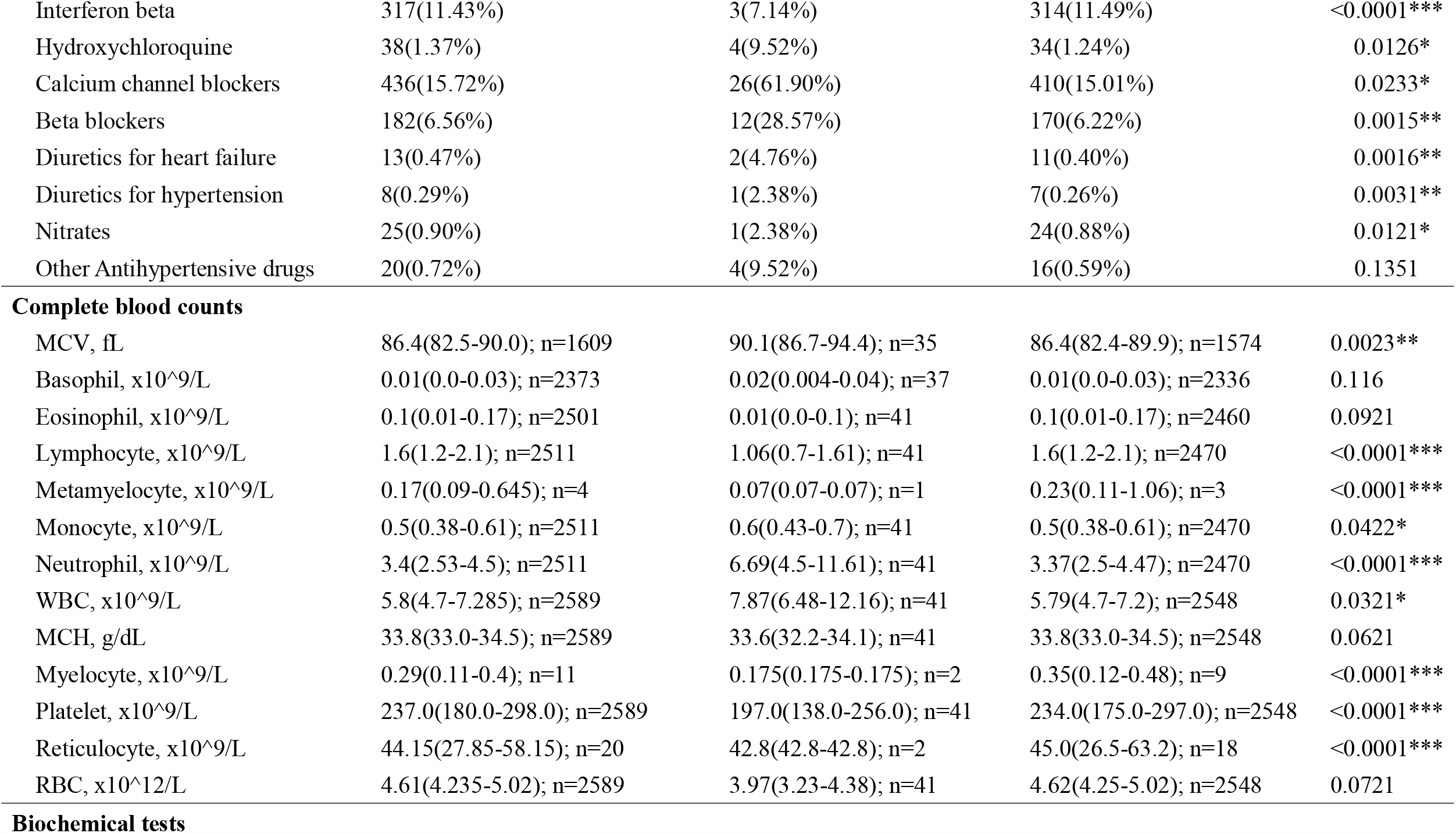
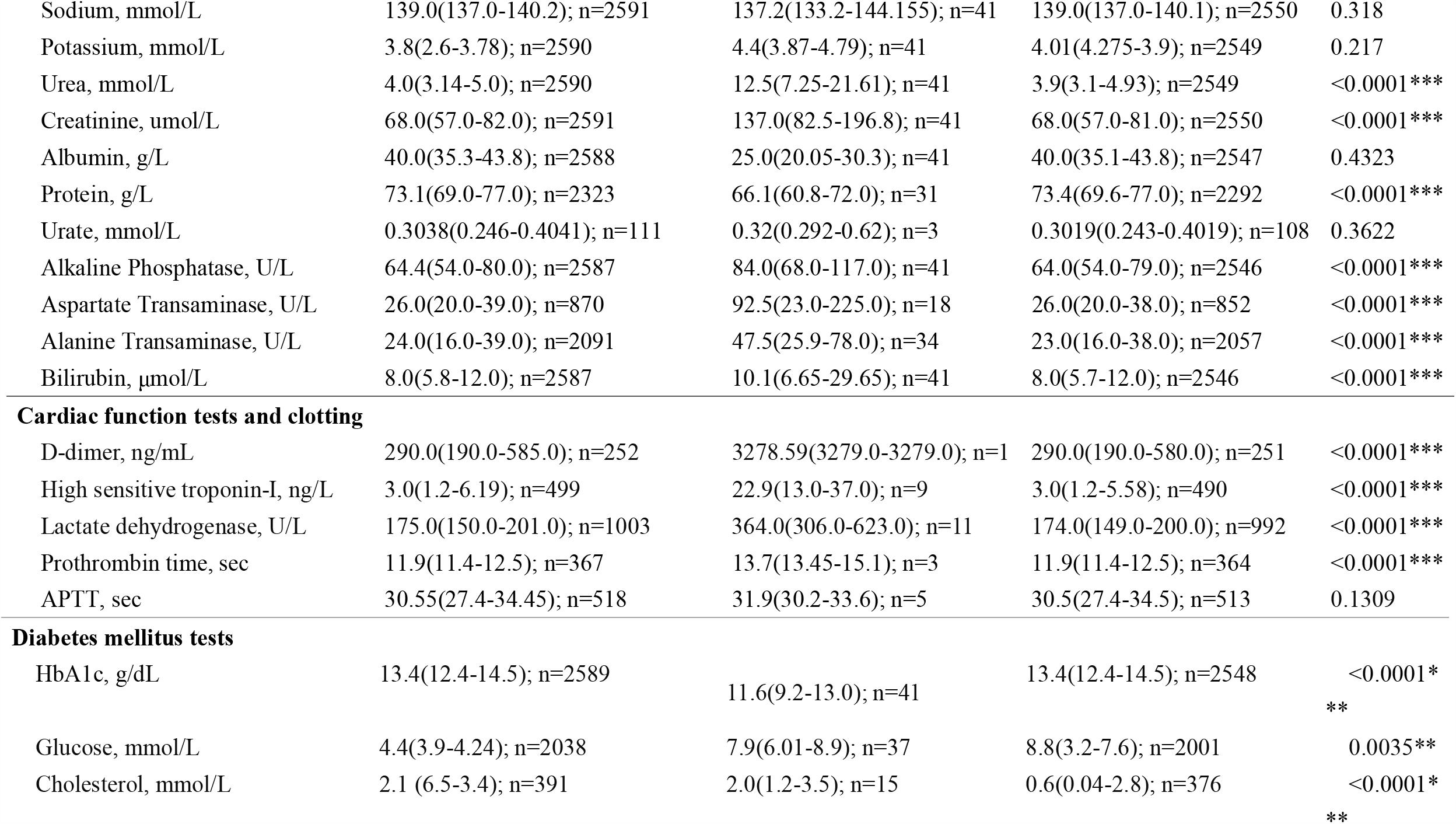
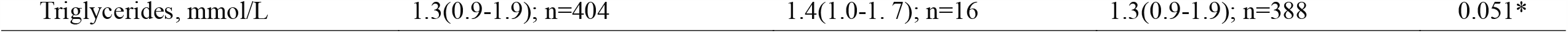
Demographic, epidemiological, clinical characteristics and laboratory details of patients who were tested positive for COVID-19 in Hong Kong up to and including 28^th^ July 2020. Patients were stratified by mortality status with latest available death information on 5^th^ August 2020. COVID-19 = coronavirus disease 2019; APTT = Activated partial thromboplastin time; IQR = Interquartile range; * for p≤ 0.05, ** for p ≤ 0.01, *** for p ≤ 0.001

| Characteristics | All COVID-19 patients (n=2774)<br>Median (IQR) or count (%) | Death (n=42)<br>Median (IQR) or count (%) | Alive (n=2732)<br>Median (IQR) or count (%) | P value |
| --- | --- | --- | --- | --- |
| **Demographics** |  |  |  |  |
| Age, year,[IQR] max | 43.54[28.61-60.00] 97.51 | 82.59[76.51,89.54] 95.59 | 42.885[28.24,59.47] 97.51 | 0.0031** |
| Sex |  |  |  |  |
| Male | 1369 (49.35%) | 26 (61.90%) | 1343 (49.16%) | <0.0001*** |
| Female | 1405 (50.65%) | 16 (38.10%) | 1389 (50.84%) | <0.0001*** |
| **Comorbidities** |  |  |  |  |
| Cardiovascular | 196 (7.07%) | 20 (47.62%) | 176 (6.44%) | <0.0001*** |
| Respiratory | 2145 (77.33%) | 36 (85.71%) | 2109 (77.20%) | 0.0122* |
| Kidney | 133 (4.79%) | 15 (35.71%) | 118 (4.32%) | <0.0001*** |
| Endocrine | 29 (1.05%) | 1 (2.38%) | 28 (1.02%) | 0.0013** |
| Diabetes | 294 (10.60%) | 18 (42.86%) | 276 (10.10%) | <0.0001*** |
| Hypertension | 413 (14.89%) | 24 (57.14%) | 389 (14.24%) | <0.0001*** |
| Gastrointestinal | 2127 (76.68%) | 32 (76.19%) | 2095 (76.68%) | 0.212 |
| **Drugs** |  |  |  |  |
| ACEI/ARB (baseline) | 279 (10.06%) | 16 (38.10%) | 263 (9.63%) | <0.0001*** |
| Steroids | 143(5.16%) | 6(14.29%) | 137(5.01%) | <0.0001*** |
| Lopinavir/Ritonavir (Kaletra) | 29(1.05%) | 4(9.52%) | 25(0.92%) | 0.355 |
| Ribavirin | 519(18.71%) | 2(4.76%) | 517(18.92%) | <0.0001*** |
| Interferon beta | 317(11.43%) | 3(7.14%) | 314(11.49%) | <0.0001*** |
| Hydroxychloroquine | 38(1.37%) | 4(9.52%) | 34(1.24%) | 0.0126* |
| Calcium channel blockers | 436(15.72%) | 26(61.90%) | 410(15.01%) | 0.0233* |
| Beta blockers | 182(6.56%) | 12(28.57%) | 170(6.22%) | 0.0015** |
| Diuretics for heart failure | 13(0.47%) | 2(4.76%) | 11(0.40%) | 0.0016** |
| Diuretics for hypertension | 8(0.29%) | 1(2.38%) | 7(0.26%) | 0.0031** |
| Nitrates | 25(0.90%) | 1(2.38%) | 24(0.88%) | 0.0121* |
| Other Antihypertensive drugs | 20(0.72%) | 4(9.52%) | 16(0.59%) | 0.1351 |
| Complete blood counts |  |  |  |  |
| MCV, fL | 86.4(82.5-90.0); n=1609 | 90.1(86.7-94.4); n=35 | 86.4(82.4-89.9); n=1574 | 0.0023** |
| Basophil, x10<sup>9</sup>/L | 0.01(0.0-0.03); n=2373 | 0.02(0.004-0.04); n=37 | 0.01(0.0-0.03); n=2336 | 0.116 |
| Eosinophil, x10<sup>9</sup>/L | 0.1(0.01-0.17); n=2501 | 0.01(0.0-0.1); n=41 | 0.1(0.01-0.17); n=2460 | 0.0921 |
| Lymphocyte, x10<sup>9</sup>/L | 1.6(1.2-2.1); n=2511 | 1.06(0.7-1.61); n=41 | 1.6(1.2-2.1); n=2470 | <0.0001*** |
| Metamyelocyte, x10<sup>9</sup>/L | 0.17(0.09-0.645); n=4 | 0.07(0.07-0.07); n=1 | 0.23(0.11-1.06); n=3 | <0.0001*** |
| Monocyte, x10<sup>9</sup>/L | 0.5(0.38-0.61); n=2511 | 0.6(0.43-0.7); n=41 | 0.5(0.38-0.61); n=2470 | 0.0422* |
| Neutrophil, x10<sup>9</sup>/L | 3.4(2.53-4.5); n=2511 | 6.69(4.5-11.61); n=41 | 3.37(2.5-4.47); n=2470 | <0.0001*** |
| WBC, x10<sup>9</sup>/L | 5.8(4.7-7.285); n=2589 | 7.87(6.48-12.16); n=41 | 5.79(4.7-7.2); n=2548 | 0.0321* |
| MCH, g/dL | 33.8(33.0-34.5); n=2589 | 33.6(32.2-34.1); n=41 | 33.8(33.0-34.5); n=2548 | 0.0621 |
| Myelocyte, x10<sup>9</sup>/L | 0.29(0.11-0.4); n=11 | 0.175(0.175-0.175); n=2 | 0.35(0.12-0.48); n=9 | <0.0001*** |
| Platelet, x10<sup>9</sup>/L | 237.0(180.0-298.0); n=2589 | 197.0(138.0-256.0); n=41 | 234.0(175.0-297.0); n=2548 | <0.0001*** |
| Reticulocyte, x10<sup>9</sup>/L | 44.15(27.85-58.15); n=20 | 42.8(42.8-42.8); n=2 | 45.0(26.5-63.2); n=18 | <0.0001*** |
| RBC, x10<sup>12</sup>/L | 4.61(4.235-5.02); n=2589 | 3.97(3.23-4.38); n=41 | 4.62(4.25-5.02); n=2548 | 0.0721 |
| Biochemical tests |  |  |  |  |
| Sodium, mmol/L | 139.0(137.0-140.2); n=2591 | 137.2(133.2-144.155); n=41 | 139.0(137.0-140.1); n=2550 | 0.318 |
| Potassium, mmol/L | 3.8(2.6-3.78); n=2590 | 4.4(3.87-4.79); n=41 | 4.01(4.275-3.9); n=2549 | 0.217 |
| Urea, mmol/L | 4.0(3.14-5.0); n=2590 | 12.5(7.25-21.61); n=41 | 3.9(3.1-4.93); n=2549 | <0.0001*** |
| Creatinine, umol/L | 68.0(57.0-82.0); n=2591 | 137.0(82.5-196.8); n=41 | 68.0(57.0-81.0); n=2550 | <0.0001*** |
| Albumin, g/L | 40.0(35.3-43.8); n=2588 | 25.0(20.05-30.3); n=41 | 40.0(35.1-43.8); n=2547 | 0.4323 |
| Protein, g/L | 73.1(69.0-77.0); n=2323 | 66.1(60.8-72.0); n=31 | 73.4(69.6-77.0); n=2292 | <0.0001*** |
| Urate, mmol/L | 0.3038(0.246-0.4041); n=111 | 0.32(0.292-0.62); n=3 | 0.3019(0.243-0.4019); n=108 | 0.3622 |
| Alkaline Phosphatase, U/L | 64.4(54.0-80.0); n=2587 | 84.0(68.0-117.0); n=41 | 64.0(54.0-79.0); n=2546 | <0.0001*** |
| Aspartate Transaminase, U/L | 26.0(20.0-39.0); n=870 | 92.5(23.0-225.0); n=18 | 26.0(20.0-38.0); n=852 | <0.0001*** |
| Alanine Transaminase, U/L | 24.0(16.0-39.0); n=2091 | 47.5(25.9-78.0); n=34 | 23.0(16.0-38.0); n=2057 | <0.0001*** |
| Bilirubin, µmol/L | 8.0(5.8-12.0); n=2587 | 10.1(6.65-29.65); n=41 | 8.0(5.7-12.0); n=2546 | <0.0001*** |
| **Cardiac function tests and clotting** |  |  |  |  |
| D-dimer, ng/mL | 290.0(190.0-585.0); n=252 | 3278.59(3279.0-3279.0); n=1 | 290.0(190.0-580.0); n=251 | <0.0001*** |
| High sensitive troponin-I, ng/L | 3.0(1.2-6.19); n=499 | 22.9(13.0-37.0); n=9 | 3.0(1.2-5.58); n=490 | <0.0001*** |
| Lactate dehydrogenase, U/L | 175.0(150.0-201.0); n=1003 | 364.0(306.0-623.0); n=11 | 174.0(149.0-200.0); n=992 | <0.0001*** |
| Prothrombin time, sec | 11.9(11.4-12.5); n=367 | 13.7(13.45-15.1); n=3 | 11.9(11.4-12.5); n=364 | <0.0001*** |
| APTT, sec | 30.55(27.4-34.45); n=518 | 31.9(30.2-33.6); n=5 | 30.5(27.4-34.5); n=513 | 0.1309 |
| **Diabetes mellitus tests** |  |  |  |  |
| HbA1c, g/dL | 13.4(12.4-14.5); n=2589 | 11.6(9.2-13.0); n=41 | 13.4(12.4-14.5); n=2548 | <0.0001*<br>** |
| Glucose, mmol/L | 4.4(3.9-4.24); n=2038 | 7.9(6.01-8.9); n=37 | 8.8(3.2-7.6); n=2001 | 0.0035** |
| Cholesterol, mmol/L | 2.1 (6.5-3.4); n=391 | 2.0(1.2-3.5); n=15 | 0.6(0.04-2.8); n=376 | <0.0001*<br>** |
| Triglycerides, mmol/L | 1.3(0.9-1.9); n=404 | 1.4(1.0-1.7); n=16 | 1.3(0.9-1.9); n=388 | 0.051* |

Univariate Cox regression was used to identify predictors of mortality in patients who were positive for COVID-19 (**Table 2**). This identified age, baseline cardiovascular and renal diseases, diabetes, hypertension, the use of ACEIs/ARBs, other anti-hypertensive agents, and anti-viral drugs in COVID-19, as well as various haematology and biochemical tests. On multivariate analysis, age, cardiovascular disease, renal disease, diabetes mellitus, the use of ACEIs/ARBs and diuretics, and various laboratory tests remained significant predictors of mortality (**Table 3**).

**Table 2.**
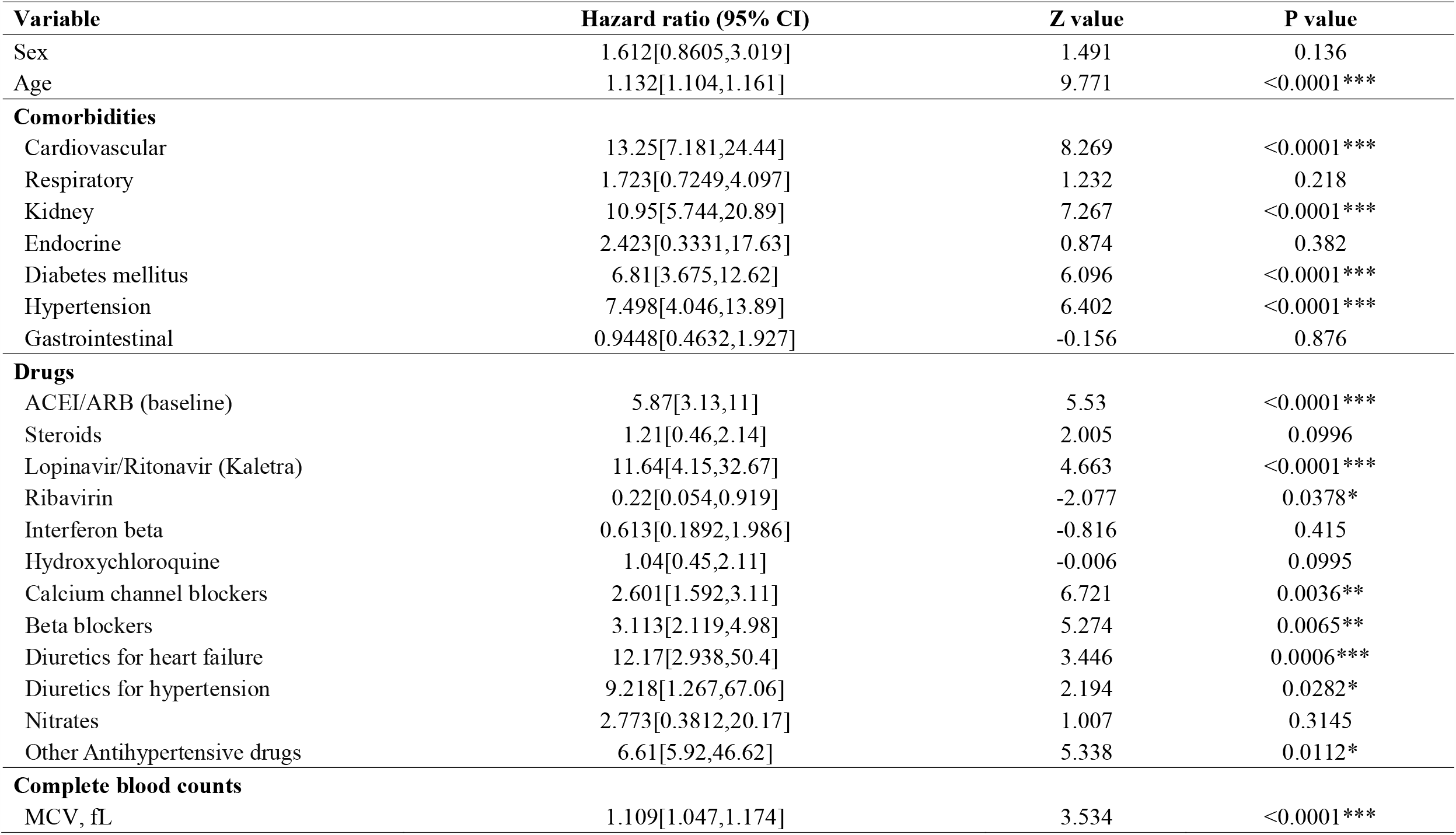
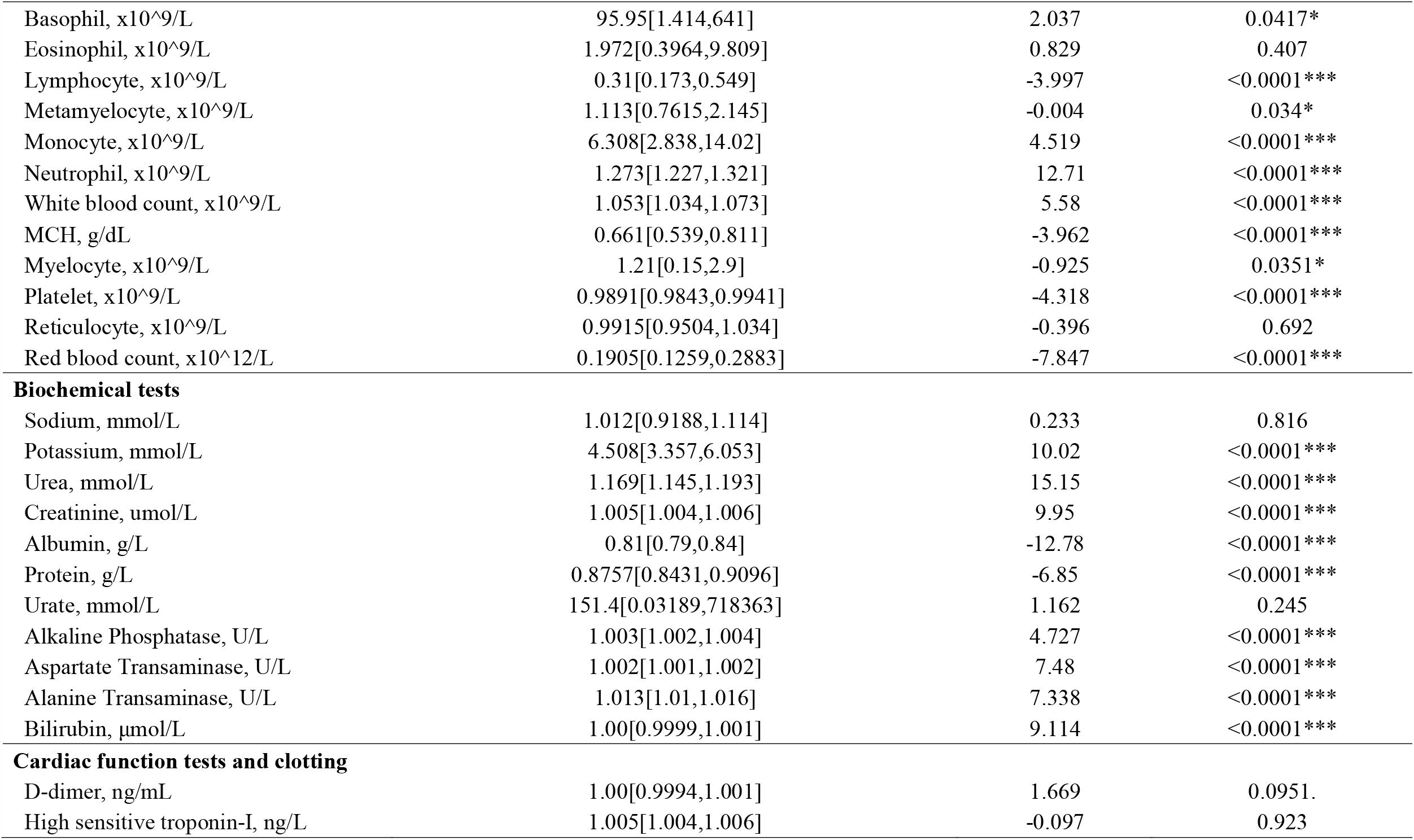
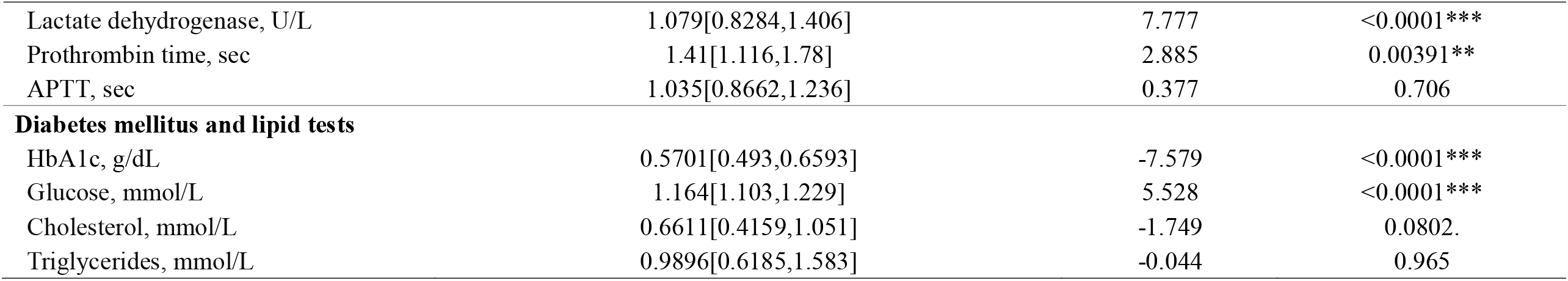
Univariate Cox regression to predict mortality after COVID-19 presentation (n=2774)

| Variable | Hazard ratio (95% CI) | Z value | P value |
| --- | --- | --- | --- |
| Sex | 1.612[0.8605,3.019] | 1.491 | 0.136 |
| Age | 1.132[1.104,1.161] | 9.771 | <0.0001*** |
| **Comorbidities** |  |  |  |
| Cardiovascular | 13.25[7.181,24.44] | 8.269 | <0.0001*** |
| Respiratory | 1.723[0.7249,4.097] | 1.232 | 0.218 |
| Kidney | 10.95[5.744,20.89] | 7.267 | <0.0001*** |
| Endocrine | 2.423[0.3331,17.63] | 0.874 | 0.382 |
| Diabetes mellitus | 6.81[3.675,12.62] | 6.096 | <0.0001*** |
| Hypertension | 7.498[4.046,13.89] | 6.402 | <0.0001*** |
| Gastrointestinal | 0.9448[0.4632,1.927] | -0.156 | 0.876 |
| **Drugs** |  |  |  |
| ACEI/ARB (baseline) | 5.87[3.13,11] | 5.53 | <0.0001*** |
| Steroids | 1.21[0.46,2.14] | 2.005 | 0.0996 |
| Lopinavir/Ritonavir (Kaletra) | 11.64[4.15,32.67] | 4.663 | <0.0001*** |
| Ribavirin | 0.22[0.054,0.919] | -2.077 | 0.0378* |
| Interferon beta | 0.613[0.1892,1.986] | -0.816 | 0.415 |
| Hydroxychloroquine | 1.04[0.45,2.11] | -0.006 | 0.0995 |
| Calcium channel blockers | 2.601[1.592,3.11] | 6.721 | 0.0036** |
| Beta blockers | 3.113[2.119,4.98] | 5.274 | 0.0065** |
| Diuretics for heart failure | 12.17[2.938,50.4] | 3.446 | 0.0006*** |
| Diuretics for hypertension | 9.218[1.267,67.06] | 2.194 | 0.0282* |
| Nitrates | 2.773[0.3812,20.17] | 1.007 | 0.3145 |
| Other Antihypertensive drugs | 6.61[5.92,46.62] | 5.338 | 0.0112* |
| **Complete blood counts** |  |  |  |
| MCV, fL | 1.109[1.047,1.174] | 3.534 | <0.0001*** |
| Basophil, x10<sup>9</sup>/L | 95.95[1.414,641] | 2.037 | 0.0417* |
| Eosinophil, x10<sup>9</sup>/L | 1.972[0.3964,9.809] | 0.829 | 0.407 |
| Lymphocyte, x10<sup>9</sup>/L | 0.31[0.173,0.549] | -3.997 | <0.0001*** |
| Metamyelocyte, x10<sup>9</sup>/L | 1.113[0.7615,2.145] | -0.004 | 0.034* |
| Monocyte, x10<sup>9</sup>/L | 6.308[2.838,14.02] | 4.519 | <0.0001*** |
| Neutrophil, x10<sup>9</sup>/L | 1.273[1.227,1.321] | 12.71 | <0.0001*** |
| White blood count, x10<sup>9</sup>/L | 1.053[1.034,1.073] | 5.58 | <0.0001*** |
| MCH, g/dL | 0.661[0.539,0.811] | -3.962 | <0.0001*** |
| Myelocyte, x10<sup>9</sup>/L | 1.21[0.15,2.9] | -0.925 | 0.0351* |
| Platelet, x10<sup>9</sup>/L | 0.9891[0.9843,0.9941] | -4.318 | <0.0001*** |
| Reticulocyte, x10<sup>9</sup>/L | 0.9915[0.9504,1.034] | -0.396 | 0.692 |
| Red blood count, x10<sup>12</sup>/L | 0.1905[0.1259,0.2883] | -7.847 | <0.0001*** |
| **Biochemical tests** |  |  |  |
| Sodium, mmol/L | 1.012[0.9188,1.114] | 0.233 | 0.816 |
| Potassium, mmol/L | 4.508[3.357,6.053] | 10.02 | <0.0001*** |
| Urea, mmol/L | 1.169[1.145,1.193] | 15.15 | <0.0001*** |
| Creatinine, umol/L | 1.005[1.004,1.006] | 9.95 | <0.0001*** |
| Albumin, g/L | 0.81[0.79,0.84] | -12.78 | <0.0001*** |
| Protein, g/L | 0.8757[0.8431,0.9096] | -6.85 | <0.0001*** |
| Urate, mmol/L | 151.4[0.03189,718363] | 1.162 | 0.245 |
| Alkaline Phosphatase, U/L | 1.003[1.002,1.004] | 4.727 | <0.0001*** |
| Aspartate Transaminase, U/L | 1.002[1.001,1.002] | 7.48 | <0.0001*** |
| Alanine Transaminase, U/L | 1.013[1.01,1.016] | 7.338 | <0.0001*** |
| Bilirubin, μmol/L | 1.00[0.9999,1.001] | 9.114 | <0.0001*** |
| **Cardiac function tests and clotting** |  |  |  |
| D-dimer, ng/mL | 1.00[0.9994,1.001] | 1.669 | 0.0951. |
| High sensitive troponin-I, ng/L | 1.005[1.004,1.006] | -0.097 | 0.923 |
| Lactate dehydrogenase, U/L | 1.079[0.8284,1.406] | 7.777 | <0.0001*** |
| Prothrombin time, sec | 1.41[1.116,1.78] | 2.885 | 0.00391** |
| APTT, sec | 1.035[0.8662,1.236] | 0.377 | 0.706 |
| **Diabetes mellitus and lipid tests** |  |  |  |
| HbA1c, g/dL | 0.5701[0.493,0.6593] | -7.579 | <0.0001*** |
| Glucose, mmol/L | 1.164[1.103,1.229] | 5.528 | <0.0001*** |
| Cholesterol, mmol/L | 0.6611[0.4159,1.051] | -1.749 | 0.0802. |
| Triglycerides, mmol/L | 0.9896[0.6185,1.583] | -0.044 | 0.965 |

**Table 3.**
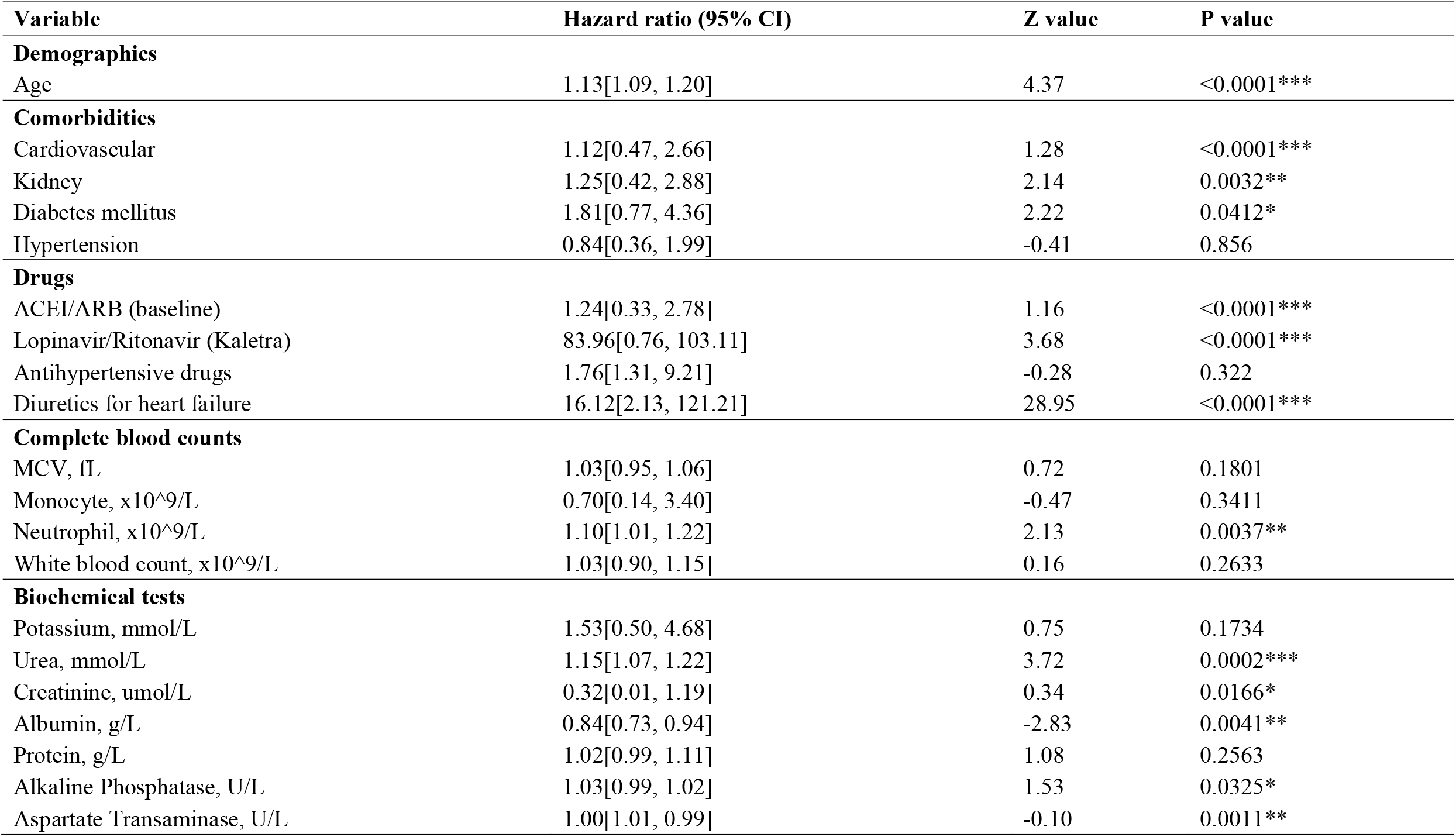
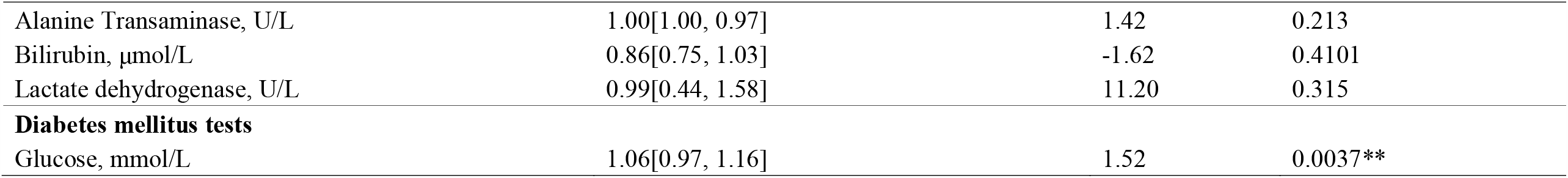
Multivariate Cox regression to predict mortality after COVID-19 presentation with significant univariate predictors as input (n=2774)

| Variable | Hazard ratio (95% CI) | Z value | P value |
| --- | --- | --- | --- |
| **Demographics** |  |  |  |
| Age | 1.13[1.09, 1.20] | 4.37 | <0.0001*** |
| **Comorbidities** |  |  |  |
| Cardiovascular | 1.12[0.47, 2.66] | 1.28 | <0.0001*** |
| Kidney | 1.25[0.42, 2.88] | 2.14 | 0.0032** |
| Diabetes mellitus | 1.81[0.77, 4.36] | 2.22 | 0.0412* |
| Hypertension | 0.84[0.36, 1.99] | -0.41 | 0.856 |
| **Drugs** |  |  |  |
| ACEI/ARB (baseline) | 1.24[0.33, 2.78] | 1.16 | <0.0001*** |
| Lopinavir/Ritonavir (Kaletra) | 83.96[0.76, 103.11] | 3.68 | <0.0001*** |
| Antihypertensive drugs | 1.76[1.31, 9.21] | -0.28 | 0.322 |
| Diuretics for heart failure | 16.12[2.13, 121.21] | 28.95 | <0.0001*** |
| **Complete blood counts** |  |  |  |
| MCV, fL | 1.03[0.95, 1.06] | 0.72 | 0.1801 |
| Monocyte, x10<sup>9</sup>/L | 0.70[0.14, 3.40] | -0.47 | 0.3411 |
| Neutrophil, x10<sup>9</sup>/L | 1.10[1.01, 1.22] | 2.13 | 0.0037** |
| White blood count, x10<sup>9</sup>/L | 1.03[0.90, 1.15] | 0.16 | 0.2633 |
| **Biochemical tests** |  |  |  |
| Potassium, mmol/L | 1.53[0.50, 4.68] | 0.75 | 0.1734 |
| Urea, mmol/L | 1.15[1.07, 1.22] | 3.72 | 0.0002*** |
| Creatinine, umol/L | 0.32[0.01, 1.19] | 0.34 | 0.0166* |
| Albumin, g/L | 0.84[0.73, 0.94] | -2.83 | 0.0041** |
| Protein, g/L | 1.02[0.99, 1.11] | 1.08 | 0.2563 |
| Alkaline Phosphatase, U/L | 1.03[0.99, 1.02] | 1.53 | 0.0325* |
| Aspartate Transaminase, U/L | 1.00[1.01, 0.99] | -0.10 | 0.0011** |
| Alanine Transaminase, U/L | 1.00[1.00, 0.97] | 1.42 | 0.213 |
| Bilirubin, µmol/L | 0.86[0.75, 1.03] | -1.62 | 0.4101 |
| Lactate dehydrogenase, U/L | 0.99[0.44, 1.58] | 11.20 | 0.315 |
| **Diabetes mellitus tests** |  |  |  |
| Glucose, mmol/L | 1.06[0.97, 1.16] | 1.52 | 0.0037** |

## Discussion

The main findings of this territory-wide cohort study are that 1) a higher proportion of ACEI/ARB prescription was observed for the COVID-19 positive compared to COVID-19 negative group and 2) those who died had a higher median age and more likely to have baseline comorbidities of cardiovascular disease, diabetes mellitus, hypertension, and chronic kidney disease and 3) ACEI/ARB use was significantly associated with mortality even after adjusting for cardiovascular comorbidities and the use of other anti-hypertensive agents. Moreover, they were more frequently prescribed ACEI/ARBs at baseline, and steroids, lopinavir/ritonavir, ribavirin and hydroxychloroquine during admission.

The relationship between ACEI/ARB use and disease outcomes in COVID-19 has been investigated. Many of the previous studies have included hospitalized patients only. For example, in a multi-centre study from China, it was found that in hospitalized patients with COVID-19 and existing hypertension, ACEI/ARB inpatient users had a lower risk of all-cause mortality compared with nonusers [12]. In our study, we found that in those who died, there was a higher frequency of patients with ACE inhibitor/ARB use compared to those who remained alive. However, on multivariate Cox regression, ACEI/ARB use retained its significance for mortality prediction even after multivariate adjustment for cardiovascular comorbidities and the use of other anti-hypertensive agents. Nevertheless, the frequency of hypertension, diabetes mellitus and cardiovascular disease were also higher in the mortality group. This is in keeping with previous demonstrations that patients with pre-existing cardio-metabolic comorbidities are at higher risk of severe disease or mortality [13].

In terms of laboratory findings, prior studies have demonstrated the presence of organ damage in COVID-19. Indeed, there have been reported associations between elevations in myocardial biomarkers and mortality [14-16]. Alterations in other laboratory markers that have been associated with adverse outcomes in COVID-19 include increases in D-dimer levels [17], liver enzymes [18], neutrophil count [19], activated partial thromboplastin time and prothrombin time [20]. In our study, we also found that patients who died had a higher white cell count, higher neutrophil count, lower platelet count, prolonged prothrombin time and activated partial thromboplastin time, D-dimer, troponin, lactate dehydrogenase, creatinine, alanine transaminase, aspartate transaminase and alkaline phosphatase.

## Conclusion

Taken together, we report that an association between ACEIs or ARBs with COVID-19 related mortality even after adjusting for cardiovascular and other comorbidities, as well as medication use. Our findings suggest that patients with greater comorbidity burden and laboratory markers reflecting deranged clotting, renal and liver function, and increased tissue inflammation, as well as ACEI/ARB use have a higher mortality risk.

## Data Availability

Anonymized data available upon request for confirmation of reproducibility.

## Contributorship Statement

GT: study conception, study design, data acquisition, statistical analysis, data interpretation, manuscript drafting, critical revision of manuscript

JZ: statistical analysis, data interpretation, manuscript drafting, critical revision of manuscript

SL, WTW, XW, WKKW, TL, ZC, DZ, KCKW: data interpretation, critical revision of manuscript

BMYC, QZ: project administration and supervision, statistical analysis, data interpretation, critical revision of manuscript

Guarantor of overall content: GT

## Acknowledgements

None

